# Using discrete Ricci curvatures to infer COVID-19 epidemic network fragility and systemic risk

**DOI:** 10.1101/2020.04.01.20047225

**Authors:** Danillo Barros de Souza, Jonatas T. S. da Cunha, Everlon Figueirôa dos Santos, Jailson B. Correia, Hernande P. da Silva, José Luiz de Lima Filho, Jones Albuquerque, Fernando A. N. Santos

## Abstract

The damage of the novel Coronavirus disease (COVID-19) is reaching unprecedented scales. There are numerous classical epidemiology models trying to quantify epidemiology metrics. Usually, to forecast epidemics, classical approaches need parameter estimations, such as the contagion rate or the basic reproduction number. Here, we propose a data-driven, parameter-free, geometric approach to access the emergence of a pandemic state by studying the Forman-Ricci and Ollivier-Ricci network curvatures. Discrete Ollivier-Ricci curvature has been used successfully to forecast risk in financial networks and we suggest that those results can provide analogous results for COVID-19 epidemic time-series. We first compute both curvatures in a toy-model of epidemic time-series with delays, which allows us to create epidemic networks. By doing so, we are able to verify that the Ollivier-Ricci and Forman-Ricci curvatures can be a parameter-free estimate for identifying a pandemic state in the simulated epidemic. On this basis, we then compute both Forman-Ricci and Ollivier-Ricci curvatures for real epidemic networks built from COVID-19 epidemic time-series available at the World Health Organization (WHO). Both curvatures allow us to detect early warning signs of the emergence of the pandemic. The advantage of our method lies in providing an early geometrical data marker for the pandemic state, regardless of parameter estimation and stochastic modelling. This work opens the possibility of using discrete geometry to study epidemic networks.

## I. INTRODUCTION

Epidemic outbreaks represent a significant concern for global health. The COVID-19 outbreak has caught the attention of researchers worldwide due to its rapid spread, high fluctuation in the incubation time and uncertain health and economic outcomes. One of the urgent challenges of this outbreak concerns the development of a coordinated and continuous data-driven feedback system that could quantify the spread and the risk of the epidemic, without strongly depending on parameter estimation (such as the contagion rate or the basic reproduction number) and even when data is heterogeneous or subject to noise. Such a data-driven system would allow to develop adequate responses at different scales (global, national or local) and to allocate limited resources in the most effective ways.

The current pandemic scenario is also challenging network science. Several papers are using network approaches to investigate spreading control, isolation policies and social distancing strategies as an attempt to provide a better understanding of the COVID-19 pandemic [1-5]. Recent developments in topological and geometric data analysis [6-10] offer useful perspectives regarding real data treatment, having yielded outstanding results over the past years across many fields [11-14]. As an emerging and promising approach in network science and complex systems more generally [15], topological and geometric data analysis describes the shape of the data by associating data with high dimensional objects [6, 13, 16].

Among the numerous successful interdisciplinary applications of applied geometry and topology, ranging from differentiating cancer networks [17] to modeling phase transitions in brain networks [18], one idea in particular can be beneficial to infer the emergence of a pandemic state from COVID-19 epidemic networks using network geometry. As this approach is completely data-driven, it could provide a geometric way to give insights into the pandemic without the need for parameter estimations. Our idea is inspired by earlier results obtained for financial networks [19], where the authors showed that it was possible to relate financial network fragility with the Ollivier-Ricci curvature of a network. Most importantly, the Ollivier-Ricci curvature emerged as a data-driven “crash hallmark” for major changes in stock markets over the past 15 years. In their study of market fragility, they used these geometric tools to analyse and characterize the interaction between the economic agents (the nodes of a financial network) and their correlation levels (which defines the edges’ weights). In addition, these tools also allowed them to track the curvature of the financial network as a function of time, i.e. how the shape of the financial network changed according to a dynamic economic scenario.

As a result, the Ollivier-Ricci curvature emerged as a strong quantitative indicator of the systemic risk in financial networks. Motivated by this result, we initially used this discrete version of Ricci curvature for our approach to epidemic networks. From an implementation perspective, however, the Forman-Ricci curvature has proved to be an alternative, simpler discretization for computing the curvature. It empirically correlates with the Ollivier-Ricci curvature even being purely combinatorial, with the added value that the Forman-Ricci curvature has a faster computation time for large-scale, real-world networks [20]. Taking all this features in consideration, here, we are going to compare our results for both Ollivier-Ricci and Forman-Ricci discretizations applied to epidemic networks.

This paper is structured as follows: In the section II, we provide the definitions of Ollivier-Ricci and Forman-Ricci curvatures, respectively, followed by our introduction of “epidemic networks”, as well as network filtration. In section III, we describe our results for epidemic COVID-19 networks using the discrete Ricci curvatures for synthetic and real data. Finally, in section IV, we present the conclusions and wider implications of our work.

## II. METHODS

This section discusses both the Olliver-Ricci and Forman-Ricci curvatures, as well as the filtration methods that we will apply applied.

### A. Ollivier-Ricci Curvature

The Ollivier-Ricci curvature for networks is defined as follows [20, 21]: Let *G* = (*V*, *E*) be a weighted undirected graph, where *V* and *E* denote its vertices and edges, respectively. The path length function *d*: *V* × *V* → ℝ^+^ defined as the length of the shortest path between two nodes induces a metric for the set of nodes of *G*. The neighborhood of a node *x* ∈ *V* is the subset of nodes connected to x by an edge, and is denoted by *π_x_*. Let *α* ∈ [0,1] and *x* ∈ *V*. We define 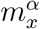, a probability measure over the set of nodes as

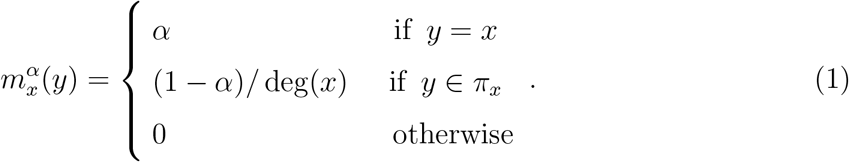

Finally, the Olliver-Ricci curvature of an edge *e* = (*x*, *y*) ∈ *E* is defined as

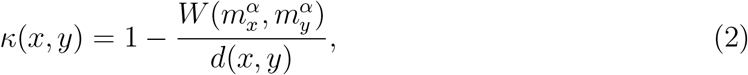

where *W* is the discrete Wasserstein distance ([9]) given by

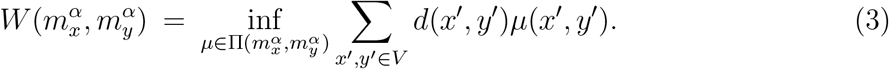

Here, 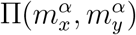 denotes the set of all probability measures *μ*: *V × V →* ℝ^+^ that satisfy

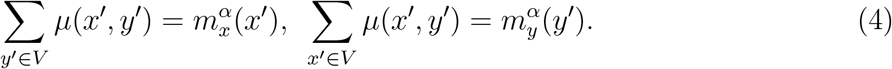

In this way, the computation of the Ollivier-Ricci curvature, in equation (2), requires solving an optimization and linear programming problem provided by equations (3) and (4), respectively. In our work, we set *α* = 0.5. Although Ollivier-Ricci discretization has numerous applications across fields [19, 22, 23], the implementation can be time consuming when compared to other discrete version of network curvatures. For this reason, we used and compared, alternatively, the combinatorial version of the Forman-Ricci curvature, as discussed below.

### B. Forman-Ricci Curvature

The Forman-Ricci curvature is simply defined as follows [20, 24]: Let *G* = (*V*, *E*) be an undirected graph. A *p*-cell in *G* is a space that is homeomorphic to an open disk of dimension *p*. As illustrated in FIG. 1, nodes are 0-cells, edges are 1-cells, triangles are 2-cells and so on. The set of all *p*-cells in a graph *G* is the *p*-skeleton of *G*. We define a CW-complex as the union of all *p*-skeletons [24, 25, 25]. We illustrate a CW-complex in FIG. 2. The Forman-Ricci curvature of a graph is calculated from its *p*-cells. Considering a CW-complex obtained from the graph and given two *p*-cells *α*_1_ and *α*_2_, we denote that *α*_1_ is contained in the boundary of *α*_2_ by *α*_1_ < *a*_2_.

**FIG. 1:**
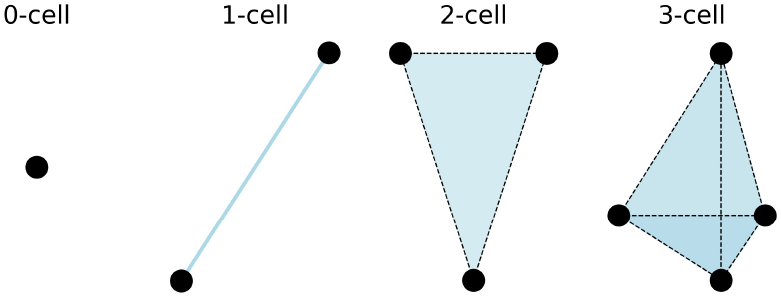
Illustration of *p*-cells of CW-complexes for *p* ∈ {0,1, 2, 3}.

**FIG. 2:**
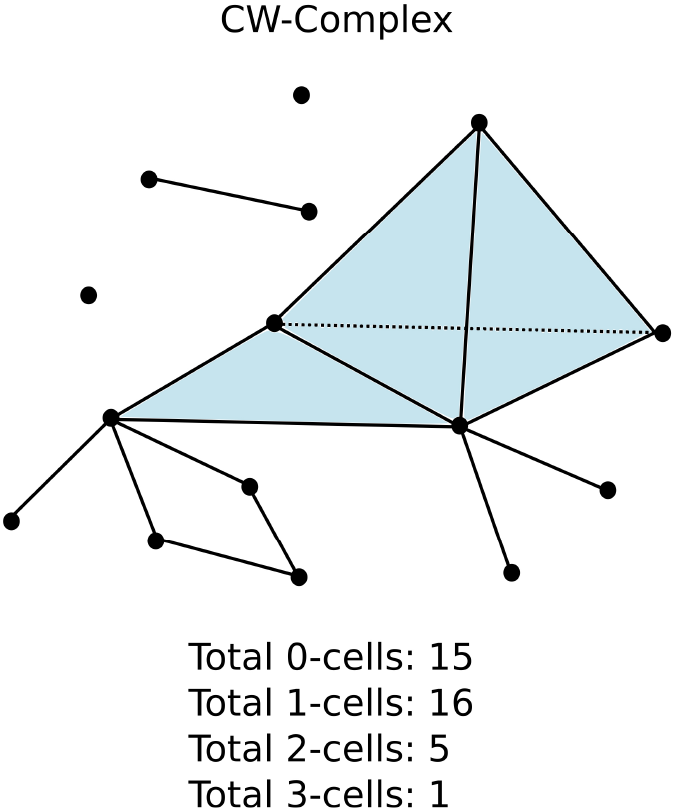
Example of a CW-complex and its *p*-cells, in this case, *p* ∈ {0,1, 2, 3}.

We say that *α*_1_ and *α*_2_ are *neighbors*, if at least one of the following conditions are fulfilled [24]:

1. There is a (*p* + 1)-cell *β* such that *α*_1_ < *β* and *α*_2_ < *β*;
2. There is a (*p* − 1)-cell *γ* such that *γ* < *α*_1_ and *γ* < *α*_2_.

We say that *α*_1_ and *α*_2_ are *parallel neighbors* if only one of the conditions 1 and 2 is true (but not simultaneously). If both 1 and 2 are true, *α*_1_ and *α*_2_ are said *transverse neighbors*. In FIG.1 we illustrate a simple example of both parallel and transverse neighbors. In FIG. 3 we illustrate the transverse and parallel neighbors of a fixed edge *e* = (*x*, *y*) in a CW-complex for *p* =1. The *p*-th Forman curvature for an unweighted *p*-cell *α* is given by [24].

**FIG. 3:**
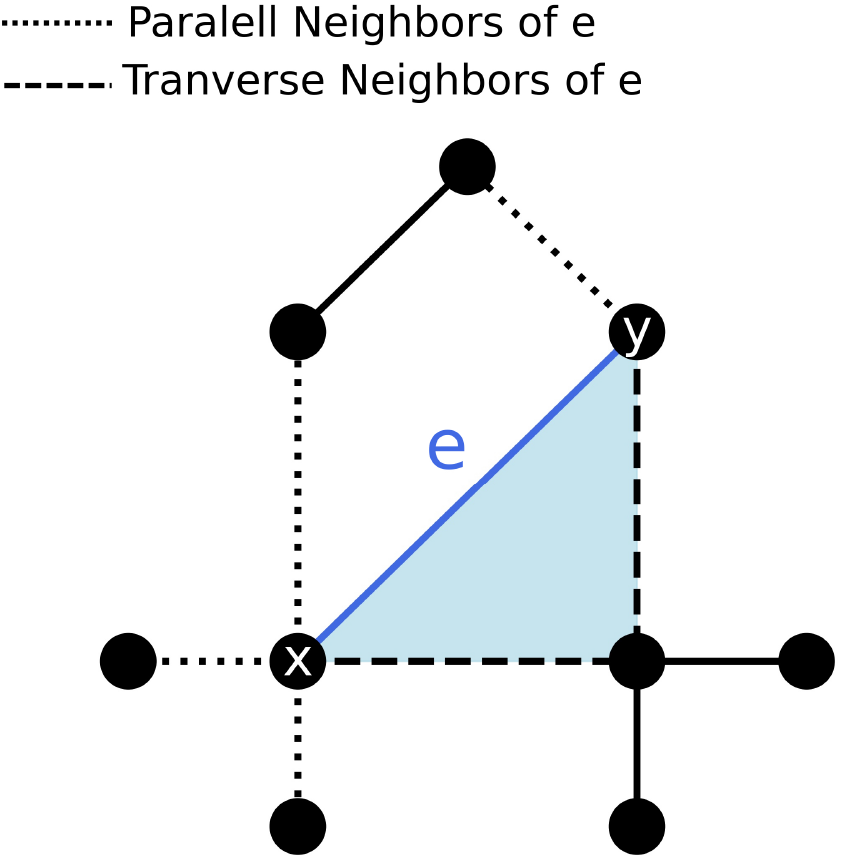
Example of parallel and transverse neighbors for an undirected graph. The nodes *x* and *y* have 5 and 3 neighbors, respectively. Following the definition, the edge *e* = (*x*, *y*) has 4 parallel neighbors (dotted edges) and 2 transverse neighbors (dashed edges).

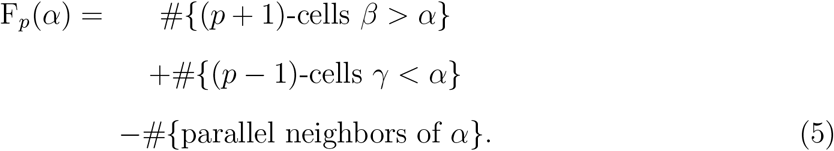

In this work we are considering vertices, edges, and triangles only, i.e., *p* =1. Therefore the Forman-Ricci curvature reduces to

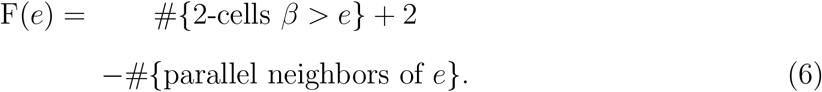

For example, in FIG. 3, the blue triangle is a (2-cell), which is bounded by the blue edge and two dashed edges (1-cells), while nodes are 0-cells. Applying (6) for *e* = (*x*, *y*), we have: one triangle (2-cell) that contains *e* = (*x*, *y*), as well as 4 parallel neighbors. Therefore, F(*e*) = 1 + 2 − 4 = −1. The average Ricci curvature is the mean of the curvatures of all edges, for both discretizations. We also extend the definition for the nodes as the mean of Ricci curvatures of the adjacent edges. In analogy with [19] and other applications of discrete versions of Ricci curvatures [17, 22, 26, 27], we provide evidence that the Forman-Ricci curvature can be used to indicate the risk and vulnerability of reaching a pandemic state.

### C. Epidemic Networks

After this illustrative explanation of the Forman-Ricci curvature, we can now introduce the concept of *epidemic networks*. In fields where network science is applicable [28], and particularly in network neuroscience [29], weighted networks are build based on correlations between nodes of a network, or any other similarity measure. Similarly, in studies of financial networks [19, 30], the resultant weighted network is built from Pearson correlations between financial time-series [30]. In contrast, in most of the network approaches to epidemiology [31, 32], the structure of the contagion network is considered, i.e., the focus is studying an epidemic in a network [32].

Here, following the network science approaches in neuroscience and economy, we introduce an *epidemic network* as a weighted network, where nodes are locations and the weights are defined by the Pearson correlation coefficient between two epidemic time-series. As far as we know, this way of defining an epidemic network from correlations was not yet introduced in the literature on network epidemiology. Therefore, not much is known regarding the relationship between our epidemic network approach and classic network epidemiology. This question deserves further investigation and can draw inspiration from other fields confronted with similar issues, in particular neuroscience [33, 34],

In this paper, we create an epidemic network consisting of edges and links, based on the reported epidemic time-series. We define each spatial domain of the epidemic as the node of a network. The links between two locations are based on the Pearson correlation coefficient (or any similarity measure) between their epidemic time-series. We chose the links of the network according to the Pearson’s correlation coefficient between two locations in descending order, which means that we include the strongest links in the network first, until the network reaches a state with a single or minimal number of clusters. Later in the paper, we discuss the sensitivity of our results to our choice of the threshold. The strategy for thresholding is based on the concept of filtration, which, for instance, [10] was used in neuroscience for classifying disease and control groups [18, 35].

More precisely, let *G* = (*V*, *E*) be a simple weighted graph. We denote the set of filtrated edges *E_ϵ_* by *G_ϵ_* = (*V*, *E_ϵ_*), where

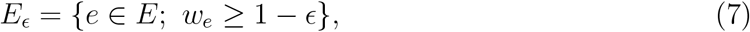

and *w_e_* is the weight of the edge *e*. Once the edges’ weights are in the interval [-1,1], ϵ runs over the interval [0, 2], and then *G*_ϵ_ ⊂ *G*. In order to ignore redundant information for the time-varying analysis, we are going to find the critical treshold value, ϵ*_c_*, such that the graph still keeps the relevant connections of the skeleton structure. This threshold value is defined as follows:

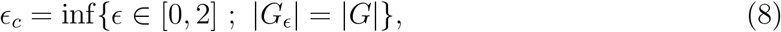

where | · | denotes the number of connected components (*β*_0_) of the graph.

## III. GEOMETRIC APPROACH TO EPIDEMIC NETWORKS

### A. Toy Model

As a proof of concept, we decided to first investigate whether the Forman-Ricci curvature is able to indicate the risk of a pandemic in a toy-model. For this, we simulated an epidemic network obtained from synthetic epidemic time-series. We will first build this simple toy-model heuristically and in a second step move towards the analysis of real COVID-19 data. A simple way to access the number of cases in an epidemic network is to use the fractal growth hypothesis, as observed in [36], where the daily number of cases n(t) in an epidemic follows a power-law distribution with an exponential cutoff:

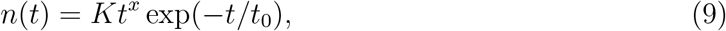

where, *K*, *x* and *t*_0_ are fitting parameters. In FIG. 4, we show examples of the fit between (9) and the number of newly reported COVID-19 cases for seven countries, namely, Brazil, China, Italy, Russia, Spain, United Kingdom, and United States. This fit suggests that (9) paves a simple way for building a toy-model for epidemic time-series. We stress that our aim here is not to find whether the best fit (or model) for the pandemic is exponential or power law, which was already addressed in [36, 37]. Instead, our goal in generating synthetic data for a pandemic is to provide a proof of concept for the applicability of both the Ollivier-Ricci and the Forman-Ricci curvatures in epidemic networks.

**FIG. 4:**
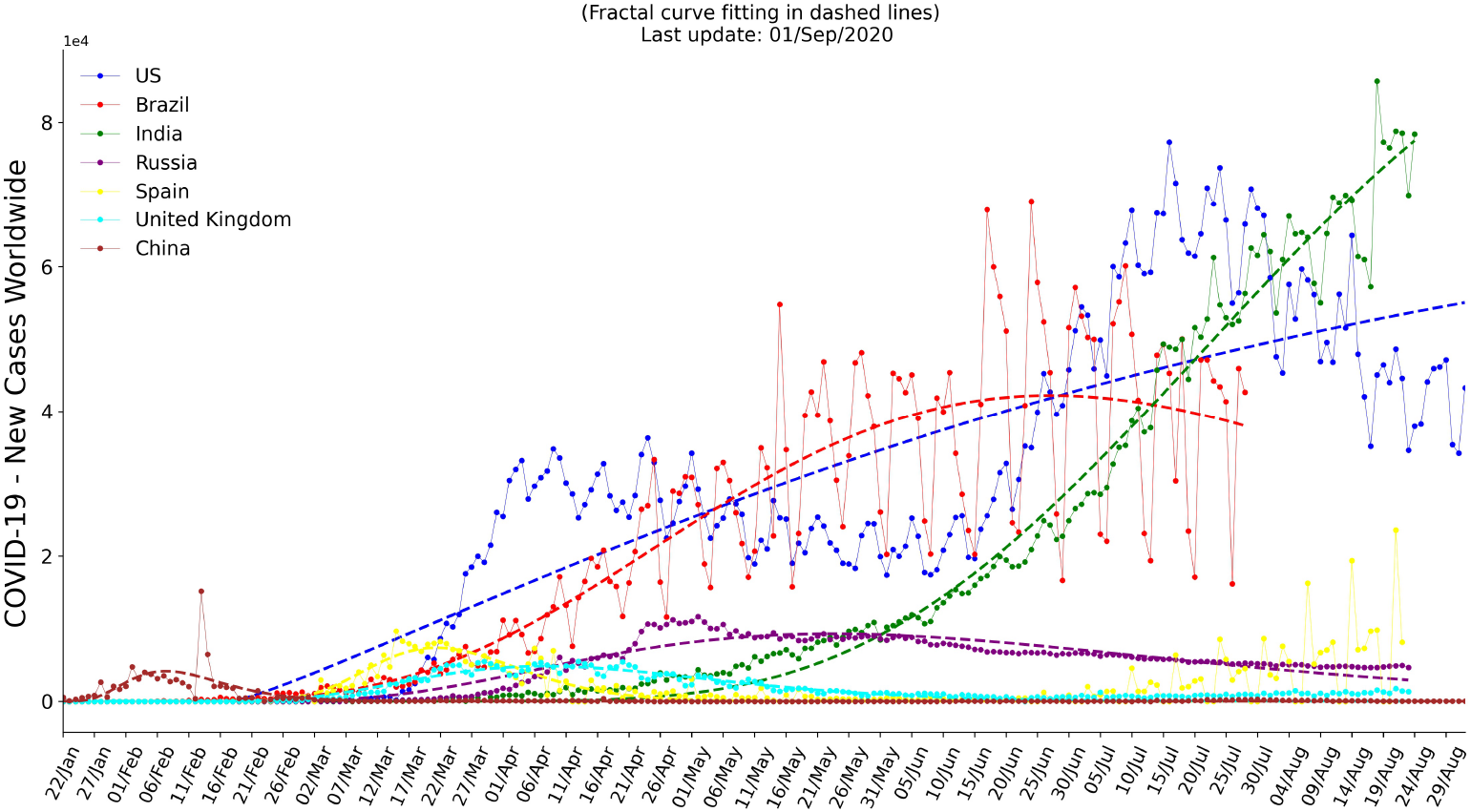
COVID-19 per country. Illustration of the daily number of newly recorded cases and fitting through fractal growth (dashed lines), Eq. (9), for a representative number of countries.

Inspired by the above equation, we can suggest a phenomenological toy-model for generating epidemic time-series with noise that can capture the growth of an epidemic network. We assume that in each node *i* of the epidemic network, the daily number of cases follows a fractal epidemic growth with Gaussian noise *w_i_* (*t*) and a time delay d_i_ in relation to the epicenter:

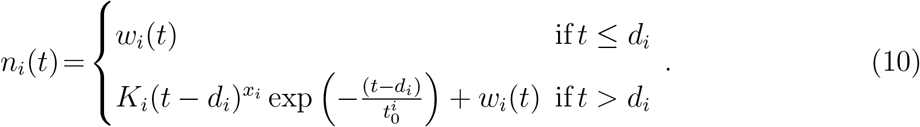

We now illustrate that we can infer the emergence of a pandemic state using both Ollivier-Ricci and Forman-Ricci curvatures. The starting point for creating a fractal epidemic network is based on simulating epidemic time-series with delays from (10). In a second step, we define the weights of the epidemic network through the Pearson correlation coefficient between time-series *n_i_*(*t*) and *n_j_*(*t*). The temporal epidemic network is computed for a given time window, and the process is repeated for the next time window, thus obtaining an evolving network. This approach is inspired by network analysis in other fields, such as neuroscience [29] or finance [30]. We illustrate the delayed epidemic time-series, its Pearson correlation matrix and its corresponding network for a given time point in FIG. 5, resulting in a time evolving network.

**FIG. 5:**
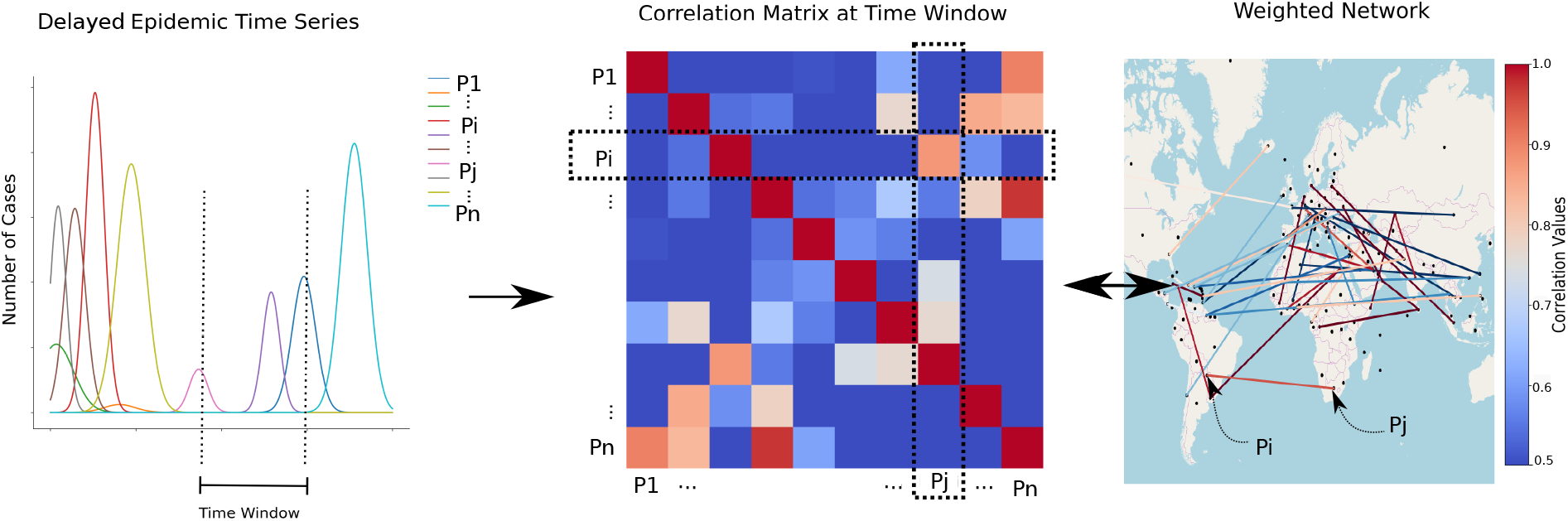
Illustration of the creation of epidemic networks based on the correlations between epidemic time-series across spatial domains for a given time window. This approach allows us to infer network signatures for epidemic outbreaks without relying on parameter estimation of classic stochastic epidemic approaches.

The third step is to infer the fragility of the time evolving epidemic network by tracking geometric changes in this network as a function of time. More specifically, we observe the mean changes in both discrete version s of the Ricci curvatures [38] for a selected moving window for each location affected by the epidemic and use the network curvature as an indicator for its fragility and risk. Given the recent results using discrete versions of the Ollivier-Ricci curvature as indicator of risk across fields, both theoretically and empirically [17, 19, 22, 26, 27], we hypothesize that the application to epidemic networks would follow similar behavior. We also implemented the Forman-Ricci curvature in an attempt to obtain an alternative, computationally efficient, geometric result.

We then investigate a simulated time-series with delays in (10). We generated 50 time-series with parameters *K_i_*, *x_i_*, *d_i_*, and 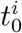 randomly chosen in the interval *K_i_ ∈* [0, 20], *x_i_* ∈ [0, 5], *d_i_* ∈ [10, 21], and 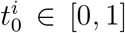. We also included a small white noise with zero mean and variance of σ = 0.01. The inclusion of white noise *w_i_*(*t*) in our toy-model was very important to destroy biased correlations that appear at the end of the outbreak (i.e., possible high correlation values that could appear between two exponentially decaying time-series). FIG. 6 shows that the epidemic curve generated from our toy-model in Eq. (10) is compatible with an epidemic outbreak and contrasts the simulated epidemic curve with both Forman-Ricci and Ollivier-Ricci curvature s.

**FIG. 6:**
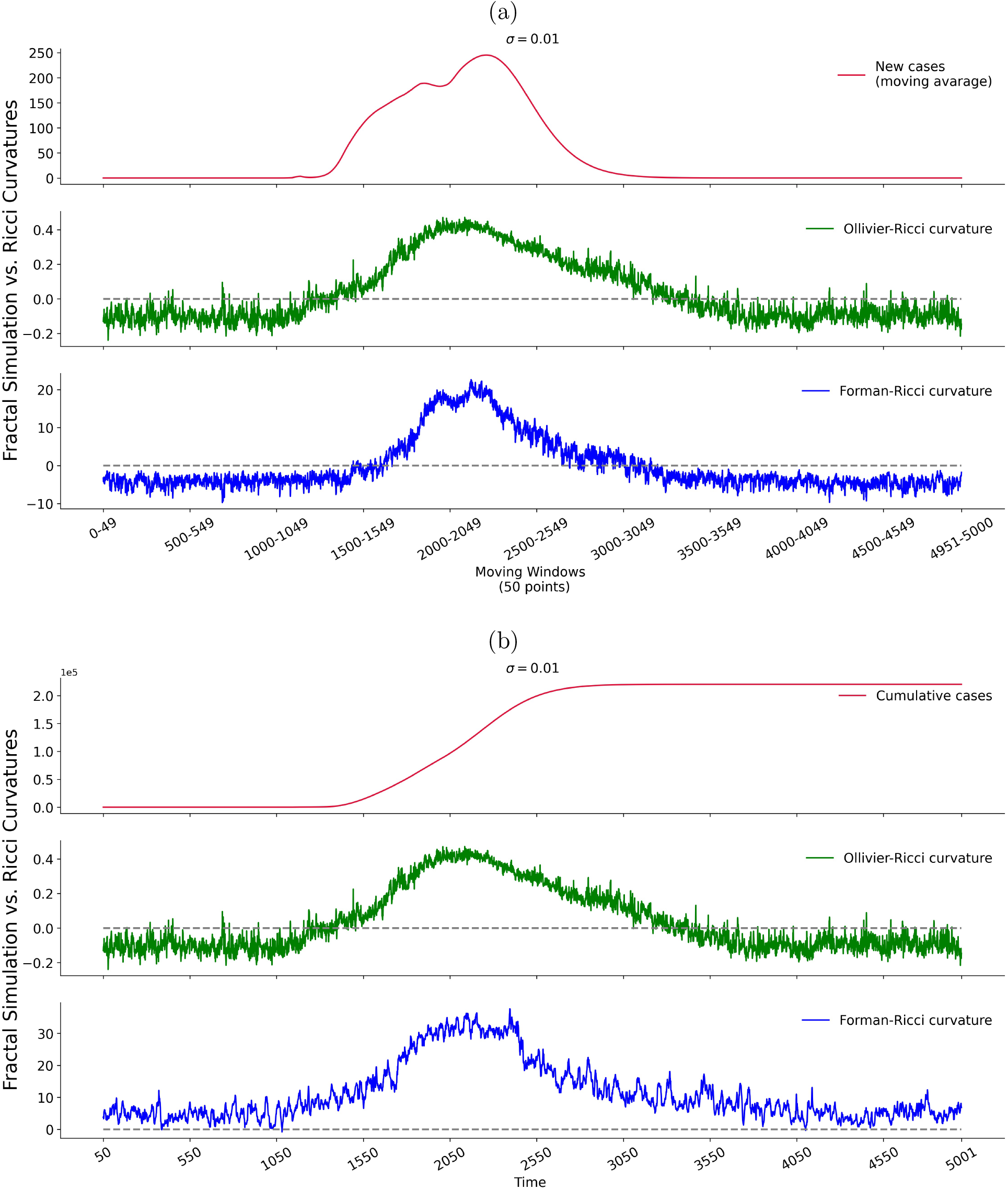
Illustration of toy-model epidemic curves, both new cases (a) and cumulative cases (b), according to (10) and its respective curvatures, with white noise parameter σ = 0.01.

We observe that the curvature is constant before the starting of the simulated epidemic, grows during its progression and reaches its maximum during the peak of the simulated outbreak. After the end of the simulated epidemic, the curvature comes back to its initial level. These results are robust when considering alternative approaches and technical choices: First, there are alternative ways for computing discrete curvatures, such as the application of Menger and Haantjes curvatures [39], as well as Ricci flow for community detection [40] and for feedback control [27]. In particular, we verified that our approach works for the Oliver-Ricci curvature and for the weighted version of the Forman-Ricci curvature. However, for rapid convergence purposes during the COVID-19 pandemics, we also decided to compare the unweighted Forman-Ricci curvature with the Ollivier-Ricci curvature. Second, we also tested the sensitiveness of our approach to different threshold values. We find that the Forman-Ricci curvature is able to identify the risk of an epidemic in both synthetic and real data for threshold values *ϵ ∈* {0.25, 0.5, 0.75,1.0,1.25,1.5,1.75}, apart from *ϵ* = 2, which leads to a fully connected graph and thus constant Forman-Ricci curvature. Third, we verified that our approach also works for different correlation metrics. We tested the Spearman and Kendall correlation coefficient instead of Pearson’s, which yielded analogous results, despite the time of processing, which may vary according to the chosen correlation coefficient.

### B. Geometric Analysis of COVID-19 Data

Having illustrated that we can use the Ricci curvatures as estimators for the risk of reaching a pandemic state in a toy-model for epidemic networks, we are now ready to test whether the discrete Ricci curvatures are a reliable network fragility measure for identifying the risk of a pandemic state for real COVID-19 data available from the World Heath Organization (WHO). In FIG. 7 we illustrate both the epidemic curve and the Ricci curvatures for the COVID-19 database [41]. The computation codes are freely available on Github [42-44]. Both discrete curvatures were based on the python code provided at [45]. We also provide Forman-Ricci computation in real time at local scales in [46].

**FIG. 7:**
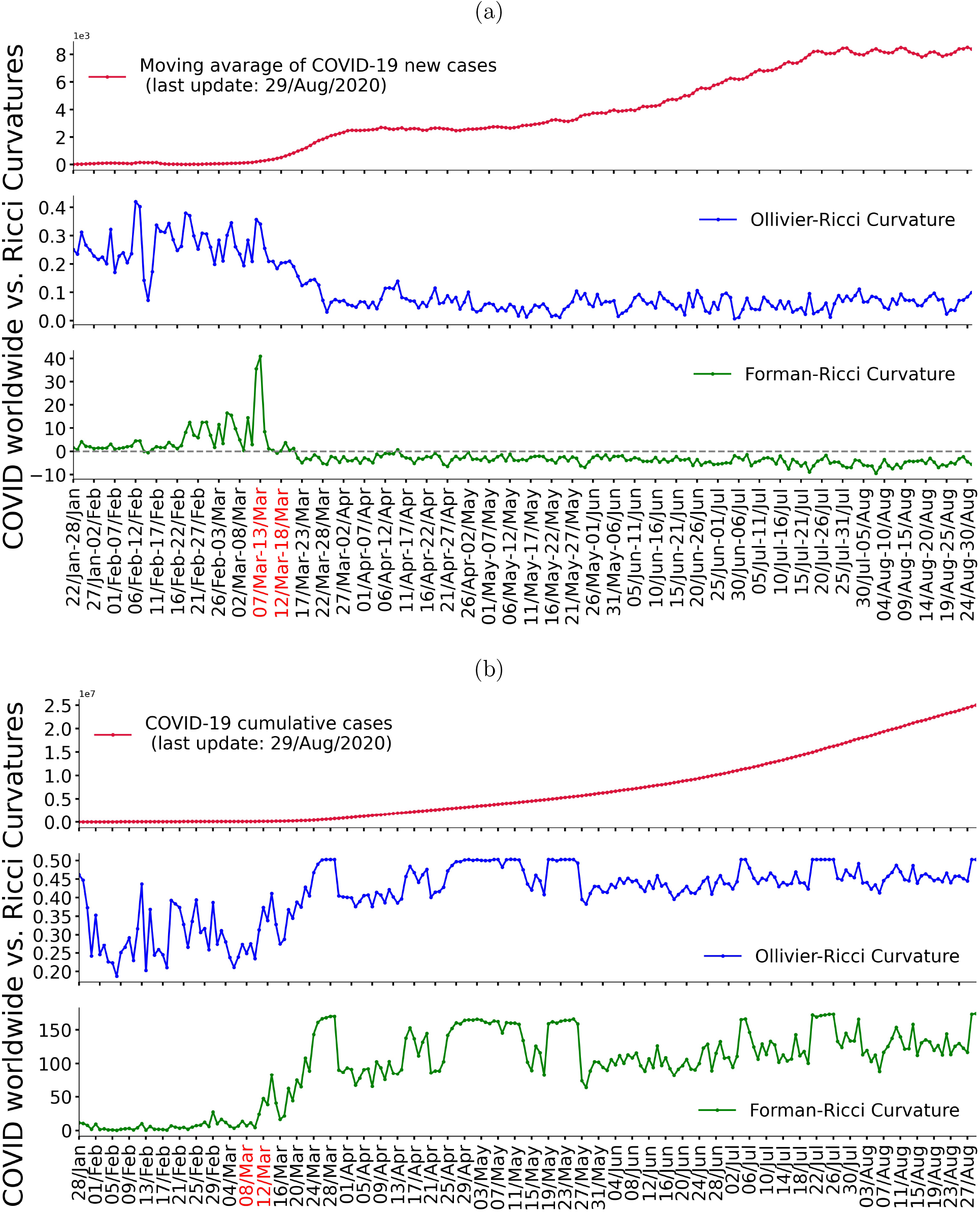
Reported epidemic cases per time window, both new cases (a) and cumulative cases (b), contrasted with its Ricci curvatures for the same time period. In red, we indicate the moment when the WHO declared COVID-19 as a pandemic. We used a time window of 7 days.

As in the simulated data, both curvatures show different geometric signatures indicating the beginning of the pandemic. Remarkably, we observe that the curvatures of the epidemic network may indicate early warning sign s for the emergence of the pandemic state, as the curvatures changes weeks before the exponential growth in number of cases is observed and the WHO declares COVID-19 as a pandemic (see FIG. 7, in red). FIG. 8 provides an additional geographical illustration of the distribution of the Forman-Ricci curvature across countries for two time windows in March 2020.

**FIG. 8:**
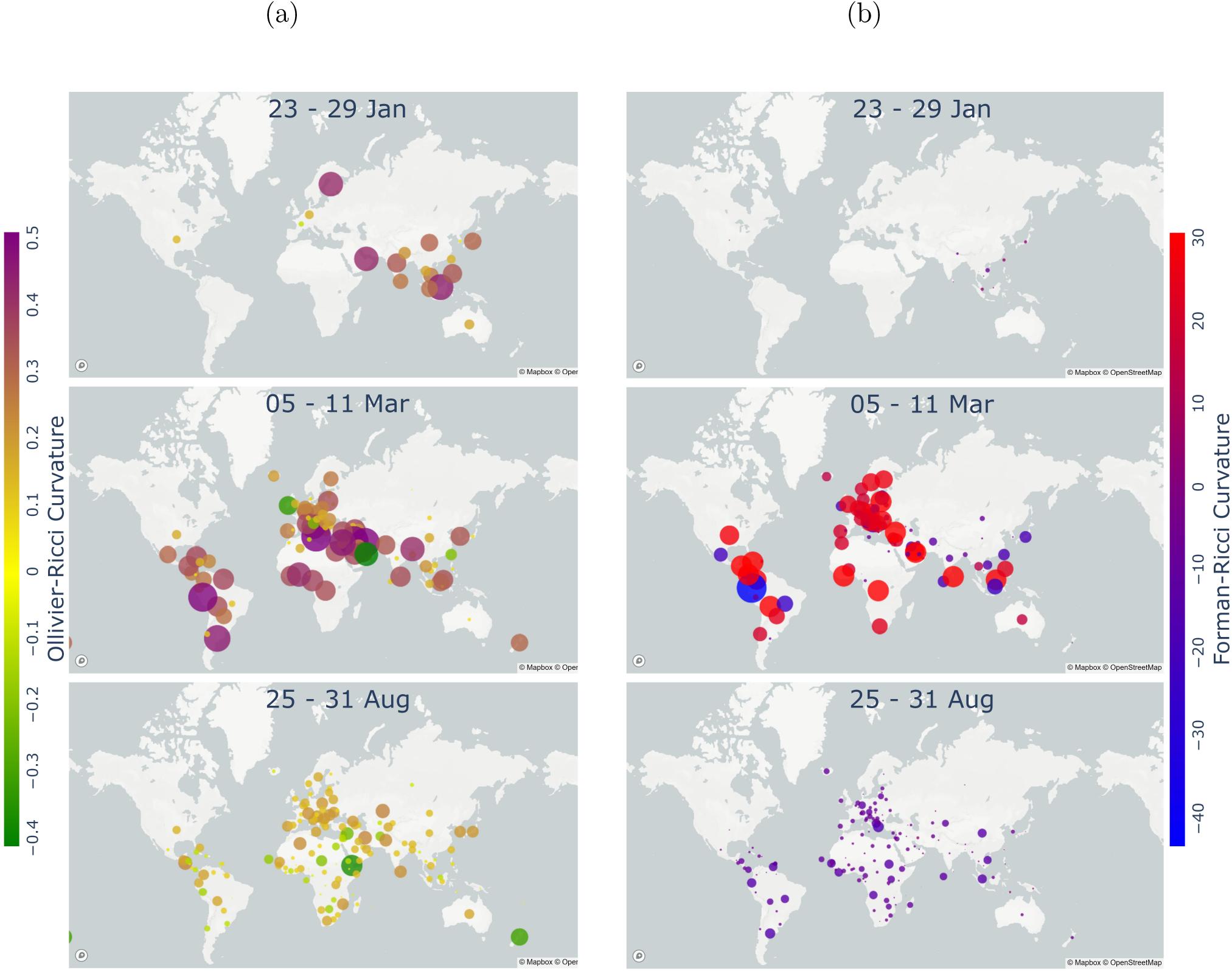
Illustration of the distribution of the Ollivier-Ricci curvature (a) and Forman-Ricci curvature (b) for three different time windows based on new cases corresponding to periods before, during, and after the beginning of the COVID-19 pandemic.

## IV. CONCLUSION

We conclude that the Ricci curvature s metric used in this paper might be a strong indicator for the identification of a pandemic state in COVID-19 epidemic networks and, consequently, could have the potential to become a data-driven, geometric approach in epidemiology more generally. Both curvatures presented similar results when it comes to detection of geometric signatures on the epidemic network. Another added value of this geometric approach, in contrast to the classical stochastic and modelling simulations, is that the results emerge intrinsically and empirically independent of parameter estimations for the pandemic. The Forman-Ricci discretization is purely combinatory and differs substantially from the Ollivier-Ricci computation. However, the Forman-Ricci curvature presents some advantages, e.g., less noise and quicker computational convergence. We tested this geometric approach for synthetic data generated by the fractal model, but the extension of this framework to other epidemic models deserves further investigation. Lastly, our work could pave the way for parameter-free and geometric approaches to epidemic networks and open the possibility for studying epidemics from a geometric perspective.

## Data Availability

The data is freely available at the World Health Organization (WHO)

## V. ACKNOWLEDGMENTS

We would like to thank critical reviews from Mariana Rossi, Katharina Natter, Dayane Torres, Silvana Bocanegra, and Fernando Moraes. This research was partially funded by INES 2.0, FACEPE grants PRONEX APQ-0602 – 1.05/14, APQ 0388 – 1.03/14 and APQ-0399 – 1.03/17, CAPES grant 88887.136410/2017 – 00, and CNPq grant 465614/2014 – 0.

D.B.S, F.A.N.S and J.A. proposed the research. F.A.N.S designed and supervised the research. D.B.S performed the research. D.B.S., J.T.S.C., and E.F.S performed the data processing and illustrations of the manuscript. D.B.S and F.A.N.S. wrote the manuscript. All authors were involved in the subsequent discussions and developments of the manuscript.

